# A Dual-Discriminator GAN and Point-Cloud Based Model for 3D Facial Appearance Prediction After Edentulous Implant Rehabilitation

**DOI:** 10.64898/2025.12.01.25341057

**Authors:** Yuwei Wu, Yangang Cai, Xiangbin Meng, Rongjie Wang, Xiangyu Yan, Chuanjie Wu, Nannan Li, Wenmin Wang

## Abstract

**Background:** Loss of all natural teeth leads to the disappearance of dental and alveolar support, causing collapse of the mid- and lower-facial soft tissues, deepening of nasolabial folds, drooping of the oral commissures, and shortening of the lower facial third. Accurately predicting postoperative facial appearance after implant-supported rehabilitation is critical for aesthetic denture design and patient communication, yet current predictions mainly rely on clinician experience and trial wax-ups.

**Methods:** We propose a point-cloud–based intelligent method and system for predicting facial appearance after edentulous implant rehabilitation. Three-dimensional facial surface point clouds of edentulous patients are acquired and combined with the 3D morphology of the planned denture. A bidirectional generative adversarial network (GAN) operating on point clouds is constructed. The forward generator *G*_*A*→*B*_ takes the edentulous face *A* and denture shape *C* as input and generates the predicted postoperative face 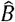; the reverse generator *G*_*B*→*A*_ reconstructs the edentulous face *Â* from the postoperative face *B*, enforcing cycle consistency. The network employs dilated convolutions with skip connections and a dual-discriminator architecture consisting of a global discriminator *D*_*G*_ for overall facial morphology and a local discriminator *D*_*L*_ focused on the perioral region. The model is trained using a weighted combination of adversarial loss, L1 loss, mean squared error (MSE) loss, and perceptual loss.

**Results:** The method was validated using pre- and post-restoration 3D facial scans from 10 edentulous patients treated with implant-supported complete dentures. Qualitative overlays showed close agreement between predicted and real postoperative faces, particularly in lip support, cheek fullness, and chin projection. Quantitatively, most surface points exhibited errors within 1.0 mm. The mean surface distance error was 0.81 mm (standard deviation 0.27 mm), and the root-mean-square error (RMSE) was 0.88 mm. Landmark displacement errors for the nasal tip, upper lip, and chin ranged from 0.5 to 1.0 mm. Predicted increases in lower-facial soft-tissue volume deviated from true values by <8%. Two experienced prosthodontists rated the agreement between predictions and outcomes with a mean visual analogue scale score of 8.5/10.

**Conclusion:** The proposed dual-discriminator point-cloud GAN achieves millimetre-level accuracy in predicting facial soft-tissue changes after edentulous implant rehabilitation without requiring CT imaging or explicit biomechanical modeling. The associated software provides real-time 3D visualization and quantitative analysis, offering an effective preoperative tool to optimize denture design and enhance patient satisfaction.

## 1. Introduction

When all teeth are lost (edentulous status), the lack of support from the dentition and alveolar bone leads to varying degrees of collapse of the mid- and lower facial soft tissues. Clinically, this manifests as deepened nasolabial folds, drooping of the oral commissures, relatively accentuated mandibular protrusion, and shortening of the lower facial third, all of which contribute to an aged appearance. Full-arch dentures, including implant-supported overdentures, can partially restore the structural support of the lower face and thereby improve facial aesthetics and proportions in edentulous patients^1,2^. However, how to quantitatively predict postoperative facial changes before implant rehabilitation remains a major challenge in aesthetic denture design^3^. At present, there is a lack of objective predictive tools in clinical practice. Treatment planning still relies largely on the clinician’s subjective judgment and experience, using intraoral wax try-ins or digital wax-ups to roughly anticipate the postoperative facial outcome^4^. These traditional approaches are time- and labour-intensive, and their efficiency is low. Moreover, patients often find it difficult to intuitively understand the design rationale and cannot preview the postoperative result in a truly digital, visual manner. Therefore, there is an urgent need for a scientific and efficient digital facial prediction method that can help clinicians foresee the aesthetic improvements prior to implant placement and optimize prosthetic design accordingly^5^.

In recent years, considerable efforts have been devoted to developing quantitative methods for predicting facial soft-tissue changes. Conventional work has mainly focused on three categories of physics-based models: (I) the mass–spring model (MSM), which discretizes the facial soft tissue into a network of mass points connected by springs to simulate deformation in real time. Although computationally efficient, MSM struggles to faithfully represent the complex mechanical behaviour of soft tissues, and its parameters are difficult to calibrate, making it prone to local over-deformation and other artefacts; (II) the finite element model (FEM), which constructs meshed models of the skull and soft tissues from CT data and simulates deformation by solving elasticity equations. FEM offers high biomechanical fidelity but is computationally expensive and time-consuming, often requiring model simplification or algorithmic acceleration to approach real-time performance; and (III) the mass tensor model (MTM), which provides a compromise between MSM and FEM by simplifying soft-tissue mechanics into a mass–tensor formulation, thereby improving computational efficiency while maintaining reasonable accuracy. For example, Mollemans et al. applied MTM to predict postoperative facial changes in maxillofacial surgery and achieved higher accuracy than MSM with substantially faster computation than FEM. Despite ongoing refinements, physics-based models generally suffer from complex model construction, challenging parameter acquisition, and limited individualization, which constrain their widespread clinical adoption^6^.

With advances in digital technology and artificial intelligence, data-driven approaches for 3D facial prediction have gained increasing attention. On the one hand, 3D facial surface scans acquired pre- and postoperatively can be combined with statistical modelling or shape analysis to quantitatively predict facial changes following dentoalveolar reconstruction^7^. For instance, Yuan et al. developed a virtual prediction system that uses a backpropagation neural network combined with a Laplacian deformation algorithm to estimate lower facial soft-tissue changes in edentulous patients wearing full dentures, achieving an average prediction error of approximately 0.77 mm. On the other hand, deep learning provides an end-to-end framework for 3D morphology prediction. Several studies have introduced generative adversarial networks to automatically complete and reconstruct facial point clouds or 3D meshes in cases of facial defects, achieving sub-millimetre mean surface distance errors in small-sample evaluations^8,9^. Collectively, these studies suggest that training intelligent models on patient-specific 3D facial data offers a promising route to high-precision prediction of postoperative facial appearance^8-11^. Building upon this foundation, the present work targets the specific scenario of implant-supported rehabilitation in edentulous patients and proposes a point cloud–based deep learning method for predicting facial changes, with the aim of improving prediction accuracy and clinical practicality without relying on complex imaging pipelines or detailed biomechanical modelling^9^.

## 2. Materials and Methods

### 2.1 Overall framework

The proposed method predicts postoperative facial morphology after prosthetic rehabilitation directly from preoperative 3D facial point cloud data. The overall pipeline consists of three stages: (I) data acquisition and preprocessing, (II) construction of a bidirectional point cloud generation network, and (III) model training and inference. First, preoperative 3D facial scans of edentulous patients are obtained to generate facial point clouds. In parallel, 3D point clouds of the designed dentition/prosthesis are created according to the planned prosthetic configuration. These two datasets are then spatially registered and fused to form a joint point cloud that encodes both the edentulous facial morphology and prosthetic information. This fused representation is used as the network input to generate the predicted post-rehabilitation facial point cloud^8,11,12^.

The network architecture is built on a generative adversarial network (GAN) framework and comprises a dilated-convolution–based generator and a dual-discriminator module with global and local branches^12-14^. The generator simultaneously receives information from the full face and the perioral region and outputs the predicted post-prosthesis facial point cloud. The discriminators assess the consistency between predicted and real postoperative data at two complementary scales: a global discriminator evaluates the entire facial contour, whereas a local discriminator focuses on the perioral aesthetic region. This design enforces a balance between accurate global morphology and faithful reconstruction of key aesthetic subregions^15,16^.

In terms of loss design, the training objective combines adversarial loss, L1 loss, mean squared error (MSE) loss, and perceptual feature loss into a weighted total loss function, thereby ensuring overall geometric accuracy while emphasizing fine facial details and visual realism^13,17,18^. In addition, a bidirectional learning strategy is adopted: the network learns the forward mapping from the edentulous face to the denture-supported face and, in parallel, a reverse mapping that reconstructs the edentulous face from the denture-supported face, enforcing a cycle-consistency constraint^17,19^. This bidirectional point cloud architecture forms a closed loop that strengthens shape consistency and geometric regularization, and it has been found to improve training stability and prediction accuracy, particularly under limited-sample conditions^20,21^.

The proposed method predicts postoperative facial morphology from preoperative 3D facial point clouds and denture morphology. The workflow comprises three main stages: (I) Data acquisition and preprocessing; (II) Construction of a bidirectional point-cloud GAN with dual discriminators; (III) Model training and prediction.

For each patient, we define:

*A*: 3D facial point cloud of the edentulous state (preoperative face);

*C*: 3D point cloud of the planned complete denture (prosthetic shape);

*B*: 3D facial point cloud after definitive prosthesis delivery (postoperative face; ground truth).

The forward generator *G*_*A*→*B*_ takes *A* and *C* as input and outputs the predicted postoperative facial point cloud 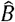:

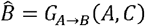

To enhance geometric consistency and training stability under limited sample sizes, a reverse generator *G*_*B*→*A*_ is introduced, reconstructing the edentulous face *Â* from the postoperative face *B*:

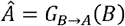

The network is thus trained to realize a cycle-consistent mapping:

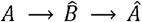

Where 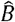 is encouraged to approximate the true postoperative face *B*, and *Â* to recover the original edentulous face *A*.

### 2.2 Data acquisition and preprocessing

A high-precision, non-contact 3D facial scanner is used to acquire dense facial surface point clouds of edentulous patients at rest, with relaxed facial muscles and gently closed lips. For a subset of patients used for validation, a second scan is obtained approximately one week after final prosthesis delivery to capture the true postoperative face *B*.

The denture morphology *C* is obtained either by scanning conventional impressions or stone casts or by importing computer-aided design (CAD) data of the planned complete denture^22,23^. Denture meshes are converted into point clouds and rigidly aligned to the edentulous facial scan of each patient.

For each case, pre- and postoperative facial point clouds are registered into a unified coordinate system using the iterative closest point (ICP) algorithm^24^. Upper-facial regions such as the forehead and orbital area are selected as reference zones because they exhibit minimal change before and after denture insertion and can be regarded as nearly rigid. In addition, anatomical landmarks such as the nasal tip, chin point, and zygomatic prominences are automatically detected and used to refine alignment of pose and scale^20,25^. This combination of landmark-based initialization and global ICP optimization effectively eliminates differences in head posture and ensures strict spatial correspondence between the pre- and postoperative faces (Figure 1).

**Figure 1.**
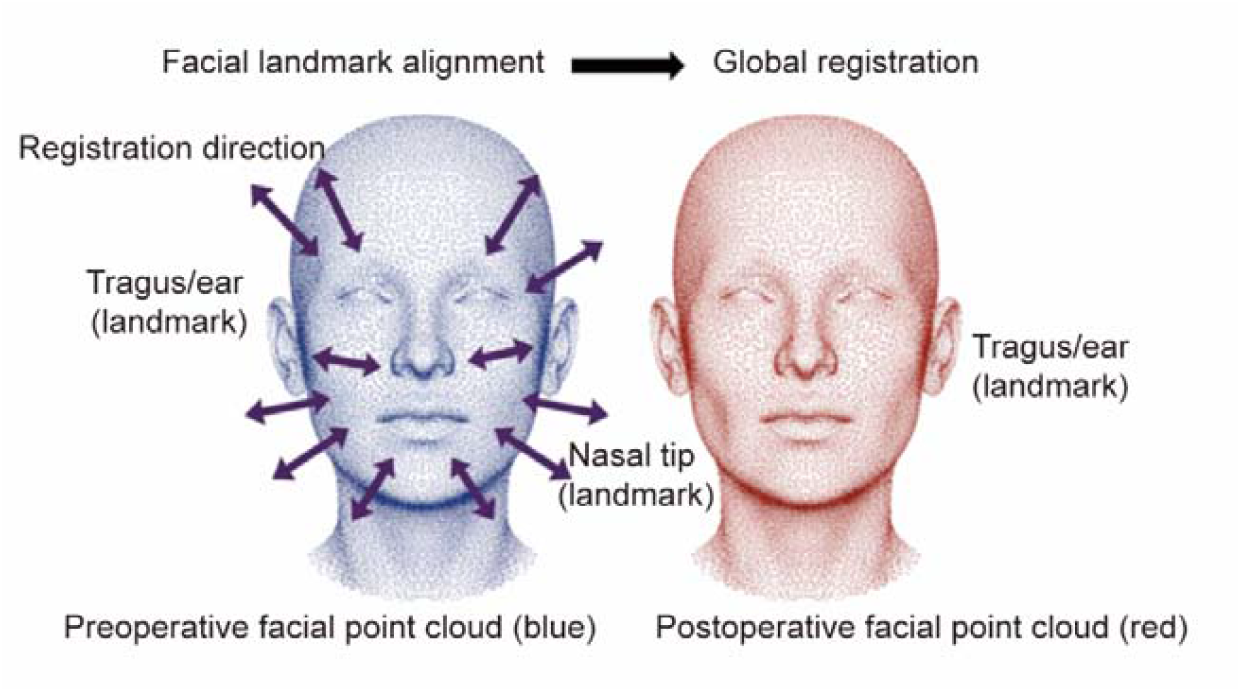
Schematic of facial landmark–based registration. Key anatomical landmarks such as the nasal tip and tragus are identified on the patient’s face to align preoperative (edentulous) and postoperative (denture-wearing) facial point clouds into a unified coordinate system, providing a consistent reference framework for subsequent prediction. The blue point cloud represents the preoperative facial surface, the red point cloud represents the postoperative facial surface, and the purple arrows indicate the registration process.

To highlight the soft-tissue changes most affected by denture support, the facial point cloud is partitioned into:(I) a global full-face point cloud, preserving the overall contour and facial proportions; (II) a local perioral point cloud, cropping the lips, cheeks, and chin area where prosthesis-induced deformations are concentrated.

The denture point cloud *C* is transformed into the same coordinate system and merged with the edentulous face *A*, providing geometric priors related to alveolar reconstruction and lip–cheek support^21,23,26^. This fused input allows the network to capture both the patient’s original facial characteristics and the volumetric contribution of the denture.

Before being fed into the network, all point clouds are normalized by translating the centroid to the origin and scaling to a common range. When necessary, principal component analysis (PCA) is applied for dimensionality reduction and noise suppression, which simplifies the model and improves robustness.

### 2.3 Bidirectional point-cloud GAN with dual discriminators

We design a deep neural network with two generators and two discriminators, forming a cycle-consistent adversarial architecture^8,12,17,19^ (Figure 2).

**Figure 2.**
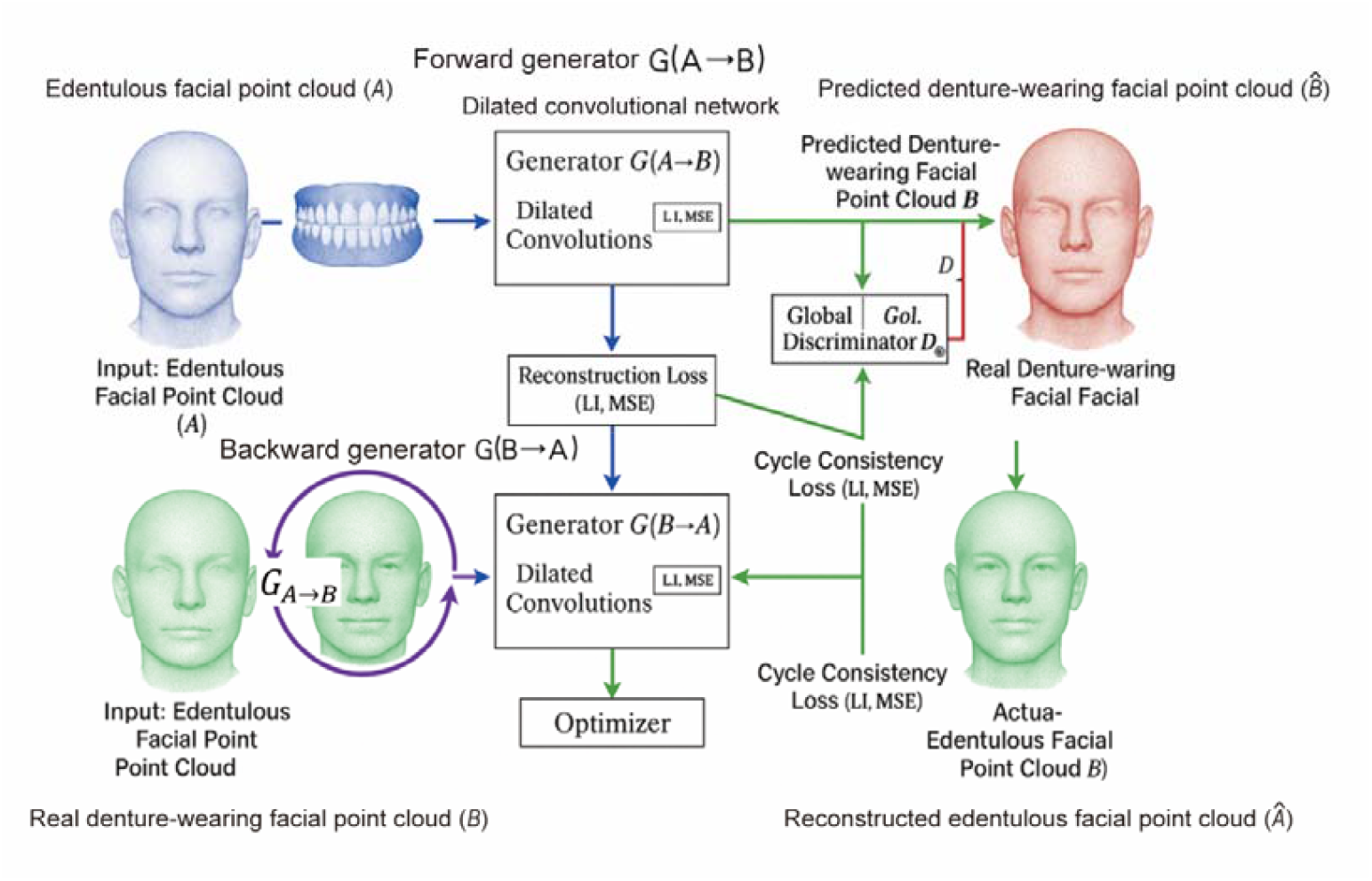
Schematic of the bidirectional point cloud generative adversarial network. The generator *G*_*A*→*B*_ takes the edentulous facial point cloud *A* and denture morphology *C* as inputs and produces the predicted denture-supported facial point cloud 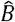. The generator *G*_*B*→*A*_ uses the real postoperative point cloud *B* to reconstruct the edentulous point cloud *Â*, providing a cycle-consistency constraint. Under the adversarial supervision of the global discriminator *D*_*G*_ and local discriminator *D*_*L*_, the predicted point cloud 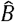 is driven to approximate the real postoperative point cloud *B*, while *Â* is enforced to remain consistent with the original edentulous point cloud *Â* Green point clouds denote generated results, red point clouds represent ground-truth data, and blue point clouds indicate input data.

The forward generator *G*_*A*→*B*_ learns the mapping from *A, C* to 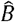.

The reverse generator *G*_*B*→*A*_ learns the mapping from *B* to *Â*

The two discriminators are:

I. Global discriminator *D*_*G*_ : Evaluates whether a full-face point cloud (either real *B* or generated 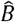) appears realistic in terms of overall shape, symmetry, and smoothness.
II. Local discriminator *D*_*L*_: Operates on a cropped perioral region and focuses on the realism of soft-tissue morphology in the lips, oral commissures, and chin, which are critical aesthetic areas^15,21^.

During training, *D*_*G*_ and *D*_*L*_ learn to distinguish real postoperative faces from generated ones, while the generators are optimized to produce predictions that fool both discriminators and satisfy reconstruction constraints.

#### 2.3.1 Loss functions

The overall objective is a weighted sum of four components: (I) adversarial loss *L*_*adv*_; (II) L1 reconstruction loss *L*_*L*1_; (III) mean squared error loss *L*_*mse*_; (IV) perceptual loss *L*_*per*_. The adversarial loss encourages the generated faces to be indistinguishable from real postoperative faces for both global and local discriminators. The reconstruction losses *L*_*L*1_ and *L*_*mse*_ measure point-wise geometric differences between generated and real faces in both the forward and reverse directions, i.e. between 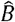 and *B*, and between *Â* and *A*. The perceptual loss *L*_*per*_ compares high-level features extracted from intermediate representations of the 3D faces, promoting visually coherent contours and smooth surfaces^13,17,18^.

The total loss is defined as:

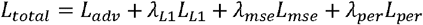

where *L*_*L*1_, *λ*_*mse*_, and *λ*_*per*_ are non-negative weighting coefficients. In this study, we empirically set

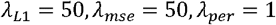

with the adversarial term normalized to unit weight, so that geometric reconstruction accuracy is emphasized while preserving perceptual quality^13,17,27^.

#### 2.3.2 Network architecture

The generators adopt an encoder–decoder structure incorporating dilated convolutions and skip connections^12,14^. Dilated convolutions enlarge the receptive field without greatly increasing the number of parameters, enabling the network to capture both local and long-range dependencies in the facial surface. Skip connections help preserve low-level geometric details and stabilize gradient flow during training.

Because point clouds are unordered sets, they are processed using point-set or graph-convolution modules inspired by PointNet and related architectures^8,26,28^. Alternatively, multi-view depth projections can be generated from the point cloud and then fed through convolutional layers. The encoded features are passed through successive convolutional and dilated convolutional blocks and finally decoded back to 3D coordinates to yield 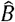 or *Â*.

The dual-discriminator design constrains the model at two scales: *D*_*G*_ enforces realistic global facial morphology, while *D*_*L*_ refines the perioral morphology. Combined with bidirectional mapping and cycle consistency, this helps the network to capture subtle protrusions and depressions induced by denture support while maintaining a natural and continuous facial envelope^11,27,29^.

### 2.4 Model training

A dataset of triplets *A*_*i*_, *C*_*i*_, *B*_*i*_ from 200 edentulous patients was used to train and validate the model. To make full use of the limited clinical data and to obtain an unbiased estimate of generalization performance, we adopted a leave-one-out cross-validation (LOOCV) strategy: in each fold, 199 cases were used for training and the remaining one case was used for validation.

During training, generators and discriminators are optimized in an alternating manner using the Adam optimizer^30^, with the learning rate set to *2* ×^-4^*10* and momentum parameters *β*_*1*_ *= 0*.*5* and *β*_*2*_ *= 0*.*999*. Training proceeds for multiple epochs until the validation error no longer decreases significantly. The final optimized model achieves high reconstruction accuracy on the validation cases, indicating that the nonlinear mapping from edentulous to postoperative faces has been effectively learned^11,27,29^.

## 3. System Implementation

To facilitate clinical use, we developed an integrated prediction system consisting of data-acquisition hardware and intelligent prediction software. The system enables fully digital workflow from facial scanning and data processing to model inference and result visualization.

### 3.1 Hardware

The hardware platform integrates a high-speed, high-precision 3D facial scanner and a workstation equipped with a dedicated graphics processing unit. The scanner can non-invasively capture high-density facial point clouds of edentulous patients in approximately one minute. For patients with existing dentures or newly designed prostheses, the system also supports importing denture models through structured-light scanning of the physical prosthesis or direct import from digital denture design software.

### 3.2 Software modules

#### 3.2.1 Dataset construction and management

This module acquires and manages all 3D data, including edentulous facial point clouds, denture point clouds, and, when available, postoperative facial point clouds^11,31^. It performs preprocessing and registration, automatically constructing a dataset for model training. The module can interface with hospital digital systems, such as electronic medical records and intraoral scanning platforms, to streamline data import and management.

#### 3.2.2 Network configuration module

This module initializes the bidirectional point-cloud GAN, including the generator and global/local discriminators, and sets hyperparameters. Pretrained model weights can be loaded directly to reduce training time and improve prediction efficiency. The architecture can also be reconfigured if new training data from different institutions are to be incorporated.

#### 3.2.3 Model training module

When new training data become available, this module conducts iterative optimization of the GAN. Training and validation losses are monitored in real time to ensure stable convergence^13,17^. After training, model parameters are frozen for clinical deployment. In typical clinical settings, users can directly employ the built-in pretrained model without on-site retraining, minimizing deployment burden.

#### 3.2.4 Facial prediction module (clinical interface)

This is the core clinical function. After importing a patient’s edentulous facial scan and denture model, the trained network generates a predicted postoperative facial point cloud within a few seconds. The result is displayed as an interactive 3D model that can be freely rotated and scaled. The system supports overlay comparison between preoperative and predicted postoperative faces, allowing clinicians and patients to jointly review changes in lip support, cheek fullness, and chin prominence. Cross-sectional analysis tools are provided to quantify displacements of key landmarks (e.g. nasal tip, labrale superius, pogonion) at millimetre precision, which assists in determining the occlusal vertical dimension and anterior tooth position^11,21^.

The system also includes report generation and data storage modules. After prediction, a standardized report containing 3D overlays, landmark displacement measurements, and cross-sectional analysis is automatically generated for clinical documentation and patient communication. All patient data and prediction records are stored in anonymized form for future model refinement while complying with data-security and privacy regulations.

Overall, the system adds only two steps to the existing workflow—an extra facial scan and a brief prediction operation—but provides clinicians with objective, quantitative, and visually intuitive preoperative facial predictions, substantially improving the scientific basis and enhancing patient understanding and satisfaction.

## 4. Experiments and Validation

### 4.1 Study population

We validated the proposed point cloud–based prediction method using prospectively collected clinical data. The study cohort comprised 10 edentulous patients (mandibular or bimaxillary edentulism) recruited from a dental hospital, aged 58–75 years (6 males, 4 females). All patients underwent full-arch implant-supported prosthetic rehabilitation and received 3D facial scanning twice: once preoperatively and once at 1 week after definitive prosthesis delivery. The high-resolution pre- and postoperative facial point clouds were preprocessed and used for method validation. For model development, data from 8 patients were used for training, and the remaining 2 patients were reserved as an independent test set to evaluate prediction performance.

### 4.2 Qualitative comparison

For each patient in the test set, the preoperative facial point cloud and the corresponding prosthesis model were input into our network to generate a predicted post-treatment facial point cloud. The predicted result was then rigidly aligned and compared with the actual postoperative facial scan of the same patient. Overlay comparisons between the predicted and actual postoperative faces showed that the predicted facial contour (green) closely matched the ground-truth contour (red dashed line), indicating accurate reconstruction of soft-tissue deformation. On visual inspection, key aesthetic changes such as anterior projection of the lips and increased fullness of the cheeks were faithfully reproduced, without obvious distortion or artefacts. These observations indicate that the method accurately captures both the direction and the magnitude of postoperative morphological changes. In contrast, without the aid of such an intelligent model, it is extremely difficult to intuitively anticipate the final appearance based solely on the preoperative facial configuration, whereas our prediction provides a clear 3D visualization that is readily understandable for both clinicians and patients.

It is worth noting that in a very small number of cases, slight postoperative asymmetry was observed clinically due to soft-tissue laxity (for example, a mildly deeper nasolabial fold on one side). Because the training data were predominantly symmetric, the model tended to predict a more symmetrical outcome in these instances. However, these discrepancies were subtle and reflected individual anatomical variation rather than systematic error, and they did not materially affect the overall qualitative assessment of prediction accuracy.

### 4.3 Quantitative error analysis

We evaluated prediction accuracy using point-to-point surface distance and volumetric overlap metrics. First, a deviation-mapping algorithm was applied to compute the distribution of surface distance errors between the predicted and reference facial point clouds for each test patient. Across the whole face, the vast majority of points showed errors within 1.0 mm, with the largest discrepancies occurring around the oral commissures, where individual points reached approximately 1.5 mm. When averaging all test cases, the mean surface distance error was 0.81 mm with a standard deviation of 0.27 mm. The root mean square error (RMSE) was 0.88 mm, indicating that the overall error magnitude is close to the intrinsic precision limit of clinical 3D facial scanning. Compared with previously reported finite element–based methods, which typically yield soft-tissue prediction errors of 2–3 mm, and with the method of Yuan et al, which reported an error of approximately 0.77 mm, our approach achieves a comparable or slightly improved accuracy, while attaining similar soft-tissue prediction performance without requiring patient CT imaging.

We further analysed the displacement of key anatomical landmarks, including the nasal tip, upper lip point, and chin point, by comparing predicted and observed vertical positional changes. On average, the prediction error for landmark displacements ranged from 0.5 to 1.0 mm. The smallest discrepancy was observed for the anterior movement of the upper lip (≈ 0.5 mm), whereas the chin point exhibited slightly larger deviations (≈ 0.9 mm). All landmark errors remained within 1.0 mm, which is considered acceptable from a clinical standpoint.

In addition, we quantified volumetric changes in the middle and lower thirds of the face. Pre- and postoperative facial point clouds were rigidly aligned in a common reference coordinate system, and the enclosed volume of the perioral and perinasal region was computed. The actual postoperative soft-tissue volume in the lower one-third of the face increased by an average of 25 cm^3^, while the model predicted an average increase of 23 cm^3^, corresponding to a deviation of less than 8%. These volumetric findings further support the effectiveness of the proposed method.

Finally, two experienced prosthodontists performed a blinded visual assessment of the predicted and actual 3D facial outcomes using a visual analogue scale (VAS; 0 = completely inconsistent, 10 = perfectly identical). The mean VAS score was 8.5, indicating that, from a clinical and intuitive perspective, the predicted facial appearance was highly consistent with the true postoperative result. Taken together, the qualitative and quantitative evidence demonstrates that the proposed method can reliably predict facial soft-tissue changes after implant-supported rehabilitation in edentulous patients and provides a valuable reference for clinical decision-making.

## 5. Results and Discussion

The point cloud–based deep learning method proposed in this study achieved robust performance for predicting postoperative facial appearance in edentulous patients. Compared with conventional approaches, it offers several distinct advantages. First, it provides high prediction accuracy. Trained on real patient data, the model is able to capture subtle trends in soft-tissue deformation, with prediction errors generally controlled within 1 mm, which is in line with the current state-of-the-art in this field. This level of precision enables prosthodontists to trust the predicted facial changes and to use them more confidently in clinical decision-making. Second, the method is highly efficient. With GPU acceleration, inference requires only a few seconds and can be seamlessly integrated into routine clinical workflows without prolonging chairside time, whereas traditional finite element simulations often take several hours and manual wax-up try-ins may require several days. Third, the approach offers strong visual interactivity. By displaying predictions on a digital 3D facial model, patients can intuitively appreciate the expected aesthetic improvements after prosthetic rehabilitation, which enhances their understanding and acceptance of the treatment plan. Such transparency has been difficult to achieve in traditional workflows and exemplifies how digital dentistry can improve patient satisfaction.

Our method, however, relies on high-quality input data and reasonable modeling assumptions. For instance, facial point cloud scans must accurately capture soft-tissue morphology. If a patient’s facial expression or head posture during scanning differs substantially from their typical facial state after wearing the prosthesis, prediction accuracy may be compromised. Therefore, during data acquisition, patients are instructed to relax facial muscles and maintain a neutral, closed-mouth position. In addition, the model assumes that the prosthesis is appropriately designed and that the patient adapts well to it. If postoperative issues such as occlusal disharmony or prolonged soft-tissue remodeling occur, the actual facial appearance may deviate from the predicted outcome. In clinical practice, model outputs should thus be regarded as an adjunctive reference, with final decisions made by clinicians based on comprehensive examination. Another important consideration is the size of the training cohort. Because fully paired pre- and postoperative datasets for completely edentulous patients are relatively limited, we adopted data augmentation and transfer learning strategies to reduce the risk of overfitting associated with small samples. The introduction of a bidirectional cyclic architecture also partly alleviates the impact of data scarcity. Nonetheless, assembling a larger and more heterogeneous dataset—particularly including edentulous patients of different ages, sexes, and craniofacial characteristics—will be crucial to further improve the model’s generalizability.

Although this study focuses on the specific clinical scenario of full-arch rehabilitation in completely edentulous patients, the proposed methodological framework is conceptually generalizable. Similar 3D point cloud–based prediction strategies could be extended to other maxillofacial reconstructive or surgical settings, such as predicting facial changes following orthognathic surgery or estimating the outcomes of soft-tissue augmentation procedures. Notably, previous studies have already applied deep learning to predict postoperative facial morphology in reconstructive and aesthetic surgery, for example in forecasting contours after mandibular defect reconstruction, and have reported encouraging results^32^. These efforts, together with our findings, collectively support the notion that data-driven 3D shape prediction has substantial potential in the maxillofacial region. In future work, we plan to strengthen collaboration with clinicians, expand multicenter clinical deployment, and continuously refine the algorithm based on real-world feedback. Furthermore, integrating multimodal information—such as CT-derived hard-tissue morphology and muscle dynamics—may further enhance the robustness and adaptability of the model across diverse soft- and hard-tissue conditions.

## 6. Conclusions

This study, based on point cloud–driven deep learning, achieves high-precision prediction of postoperative facial appearance in edentulous patients undergoing implant-supported rehabilitation. By acquiring preoperative 3D facial scans together with digital prosthesis morphology, we constructed a bidirectional generative adversarial network capable of rapidly generating the patient’s postoperative 3D facial configuration before surgery. Experimental results demonstrate that the proposed method reconstructs soft-tissue deformations induced by prosthetic support with millimetre-level accuracy, thereby addressing the long-standing reliance on clinical experience and the lack of quantitative assessment.

The intelligent prediction framework developed here can serve as a powerful adjunct to clinical decision-making. Clinicians may use it to compare alternative prosthesis designs and objectively evaluate their impact on facial aesthetics, facilitating selection of the optimal treatment plan. Patients, in turn, can visualise the anticipated postoperative outcome in advance, which helps build trust and encourages active participation in therapy. This work highlights the application potential of integrating digital dentistry with artificial intelligence and provides a novel strategy for improving aesthetic outcomes in edentulous rehabilitation. Looking ahead, with the accumulation of larger datasets and further algorithmic refinement, facial appearance prediction technology is expected to achieve even higher precision, expand its indications, and ultimately enable truly personalized, aesthetics-driven dental rehabilitation.

## Data Availability

All data produced in the present study are available upon reasonable request to the authors.

